# Continuous Reaching and Grasping with a BCI Controlled Robotic Arm in Healthy and Stroke-Affected Individuals

**DOI:** 10.1101/2025.04.16.25325551

**Authors:** Dylan Forenzo, Yisha Zhang, George F. Wittenberg, Bin He

**Author notes:** Correspondence: Bin He, Ph. D. Department of Biomedical Engineering, Carnegie Mellon University, Scott Hall 4N115, 5000 Forbes Avenue, Pittsburgh, PA, 15213.

## Abstract

Recent advancements in signal processing techniques have enabled non-invasive Brain-Computer Interfaces (BCIs) to control assistive devices, like robotic arms, directly with users’ EEG signals. However, the applications of these systems are currently limited by the low signal-to-noise ratio and spatial resolution of EEG from which brain intention is decoded. In this study, we propose a motor-imagery (MI) paradigm, inspired by the mechanisms of a computer mouse, that adds an additional “click” signal to an established 2D movement BCI paradigm. The additional output signal increases the degrees of freedom of the BCI system and may enable more complex tasks. We evaluated this paradigm using deep learning (DL) based signal processing on both healthy subjects and stroke-survivors in online BCI tasks derived from two potential applications: clicking on virtual targets and moving physical objects with a robotic arm in a continuous reach-and-grasp task. The results show that subjects were able to control both movement and clicking simultaneously to grab, move, and place up to an average of 7 cups in a 5-minute run using the robotic arm. The proposed paradigm provides an additional degree of freedom to EEG BCIs, and improves upon existing systems by enabling continuous control of reach-and-grasp tasks instead of selecting from a discrete list of predetermined actions. The tasks studied in these experiments show BCIs may be used to control computer cursors or robotic arms for complex real-world or clinical applications in the near future, potentially improving the lives of both healthy individuals and motor-impaired patients.

## I. Introduction

BRAIN-Computer Interfaces (BCIs) are devices that allow their users to control computers or other external devices directly through changes in brain activity. Non-invasive BCI systems work by recording users’ brain activity with devices like electroencephalography (EEG) and decoding intentions by signal processing. By extracting users’ intentions directly from their brains, BCIs bypass the need for muscle or speech activation and may be useful for replacing or restoring motor functions in patients with motor-impairments [1], [2], [3], [4], [5], [6], or even benefitting healthy users by providing a direct line of communication with their personal devices. An example of an EEG-BCI setup using a robotic arm is shown in Fig. 1A.

**Fig. 1.**
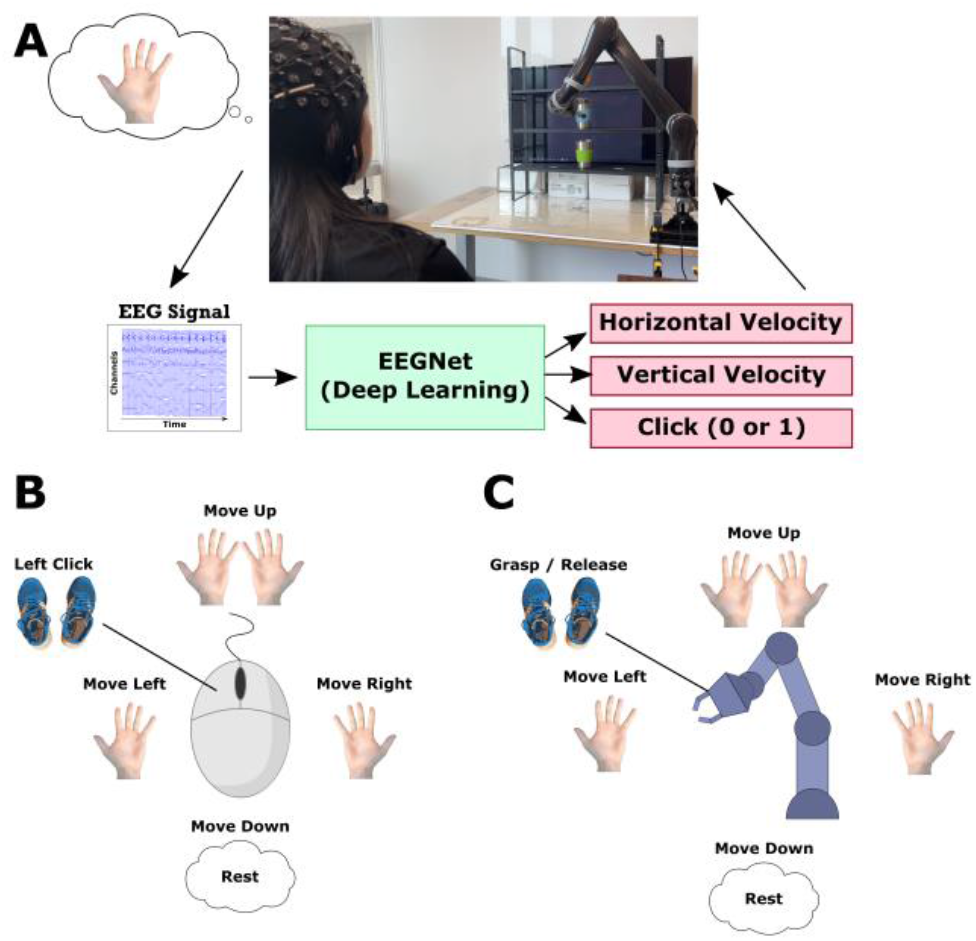
Overview of EEG-based Brain-Computer Interfaces and Motor Imagery Paradigm. A) An illustration overview of an EEG-based BCI. Starting from the top left, a human subject performs MI by imagining the movement of their left hand. Electrical signals from their scalp are then recorded by an EEG device, which can be processed using a trained DL model. The DL-based BCI decoders provide three outputs: a horizontal and a vertical velocity used to move the system in 2D, and a Boolean signal used to provide a “click”. In this example, the BCI system is used to move a robotic arm to grab and move metal cups around a vertical shelf. B) Proposed MI paradigm for virtual cursor control. Left-hand MI to move to the left, right-hand to move right, both hands to move up, rest to move down, and foot MI to click. C) Proposed MI paradigm for robotic arm control. Same 2D movement as in virtual cursor control, foot MI to initiate a grab or release.

EEG is one of the most common methods for recording BCI signals because it is non-invasive, portable, and relatively inexpensive to use [4]. Previous studies have demonstrated EEG-BCI control of a variety of devices, including: virtual cursors [7], [8], [9], wheelchairs [10], [11], [12, p. 20], [13], robotic arms [14], [15], [16], [17], [18], [19], and even a virtual or physical quadcopter drone [20], [21]. Among these applications, robotic arm control may be of particular interest since it could help to replace or restore upper limb functions in motor-impaired patients, allowing them to perform essential tasks with more autonomy.

The current state of robotic arm control using BCIs is different between invasive and non-invasive BCI methods [22]. Invasive methods can record neural signals from the vicinity of neurons with many channels simultaneously. These methods can produce high-dimensional signals with robust signal-to-noise ratios and high spatial and temporal resolutions. Using these signals, invasive BCI systems have allowed motor-impaired patients to control robotic arms to perform reaching and grasping [23], [24], move using 10 degrees of freedom, and pass clinical tests of upper-limb control [25]. More recently, invasive methods using neuro-stimulation have also added tactile feedback to robotic arm control [26], and even restored some use of patients’ own arms with the use of a functional electrical stimulation system controlled by the BCI [27]. These invasive BCIs can be a great benefit to some motor-impaired patients, but the invasive nature of their recording methods also carries a substantial risk and can limit the pool of potential users

Non-invasive BCI recording methods, like EEG, do not require any surgical procedures and are safe and cost-effective to use for the general population. However, the lower spatial resolution and signal-to-noise ratio can make it more challenging to extract users’ intentions from their EEG signals. This has limited the performance of EEG BCIs so far compared to invasive methods, but recent progress in signal processing methods has advanced the capabilities of EEG BCIs to control robotic devices. Previous studies have developed EEG BCI systems that can control robotic arms continuously in two dimensions [15], [17], [19], or perform reach and grab tasks by selecting an option from a set of predetermined actions [16], [18].

Meng et al. reported an EEG BCI controlled robotic arm, performing reaching and grasping using continuous control instead of selecting from a discrete list of choices [14, p. 20]. In this paradigm, the user controlled the robotic arm continuously in 2D to move to a target location, then initiated a grasp command by hovering in the target area for 2 seconds. This paradigm is of interest because it could be used to interact with any objects within the workspace, instead of needing to choose from a finite list of pre-set options, giving the user more control over the system. However, there is still some room for improvement in this method, since the hovering mechanism makes it difficult to determine a precious grasp location, takes additional time to send commands, and does not allow for the subject to rest, since resting would cause the cursor to hover in place and initiate a grasp.

The purpose of this study is to investigate an alternative to the hovering mechanism for continuous control of a robotic arm in reaching and grasping tasks. To accomplish this, we propose an MI paradigm that includes three output signals, two for 2D movement and an additional Boolean signal (outputs either 0 or 1), called a “click”. Using this paradigm, the BCI system can mimic a computer mouse by moving in 2D and allowing the user to *click* at will (Fig. 1B). This setup also translates to robotic arm control (Fig. 1C), allowing users to continuously control the robot in 2D and initiate a grasp or release at any location by sending a *click* signal. The proposed click paradigm has several advantages over the hovering mechanism: it allows the user to select a specific location by clicking, does not require a dwell time to select a location, and allowing the user to rest voluntarily.

The proposed paradigm has larger implications beyond just improving upon the hovering mechanism. Future BCI systems using this setup could be developed for a variety of real-world applications. In virtual tasks, the click paradigm could be used to control an actual computer mouse, allowing the BCI user to perform a variety of everyday tasks and applications on their personal computers. For physical devices, beyond the reach and grasp task, this paradigm could enable BCI systems to not only move robots in 2D, but also interact with their environment. Two examples are flying a drone and using the click signal to take pictures or driving a motorized wheelchair and using the click signal for a brake or horn. By enabling users to perform more advanced tasks, the proposed paradigm contributes to the overall goal of bringing EEG-based BCIs closer to real-world and clinical applications, potentially improving the lives of healthy individuals and motor-impaired patients.

## II. Materials and Methods

This study proposes a new motor imagery (MI) paradigm for EEG BCIs that uses MI for both moving in two dimensions and sending a Boolean *click* signal. The main experiments performed in this study included BCI sessions in which human subjects were first capped with EEG electrodes, and then asked to control the BCI system by performing MI. The subjects used the proposed BCI system to accomplish several tasks, including moving a virtual cursor to click on randomly placed targets, moving a robotic arm to virtual targets, and using the robotic arm to pick up and move physical objects around three vertical shelves. An example BCI setup of the cup task with a robotic arm is depicted in Fig. 1A.

Additional information about the study methods for training deep learning models, the traditional decoder, and robotic arm setup are provided in the Supplementary Materials.

### A. Subject Recruitment

Eight (8) healthy subjects were recruited for the study (average age: 26.125, 7 right-handed, 5 male). In addition, 5 other subjects were recruited who had a history of stroke occurring at least 3 months before participating in the study (average age: 57, 4 right-handed, 3 male). One healthy subject and one stroke survivor did not obtain a BCI performance above chance level in any of the tasks during the two BCI training sessions and were excluded from the study (50% correct in 1D tasks, 25% in 2D, see Section D. Study Design for more information). Another stroke survivor was unable to continue with the experimental sessions due to personal reasons unrelated to the study, resulting in a total of 10 subjects who completed the study (3 stroke survivors and 7 healthy subjects).

Healthy subjects were recruited through fliers placed around Carnegie Mellon University campus and the greater Pittsburgh area. Stroke-affected subjects were recruited through fliers placed in local stroke clinics and through Pitt+Me, a research subject registry operated by the University of Pittsburgh. This study was approved by an Institutional Review Board at Carnegie Mellon University (STUDY2017_00000548). All subjects were required to provide written consent to the protocol to participate. No prior BCI experience was required for participation. The healthy subjects recruited for the study had a range of prior BCI experience from no experience to having done several previous sessions with an MI BCI. All stroke-affected subjects recruited for this study had no previous experience with MI or EEG BCIs.

### B. Signal Acquisition

Subjects wore a Neuroscan Quik-Cap EEG system with 64 EEG channels amplified using SynAmps 2/RT devices from Compumedics Neuroscan. The EEG channels were arranged in a modified version of the international 10/20 system where the reference electrode is in the center of the montage, between the Cz and CPZ electrodes. All electrode impedances were reduced to below 5 kΩ before the start of each session. Digital signals were recorded using a sampling rate of 1 kHz with a low-pass filter at 200Hz and a notch filter at 60Hz to remove AC interference. The digitized EEG signals were then streamed to the BCI2000 software platform [28] in 40ms packets for downstream signal processing and cursor/robotic arm applications.

### C. Motor Imagery Paradigm

Subjects controlled the proposed BCI system using MI of their hands and feet. The proposed MI paradigm for the virtual click task and robotic arm control is illustrated in Fig. 1B and 1C, respectively. In both tasks, subjects were able to control the BCI system continuously in two-dimensions by performing left hand MI to move to the left, right hand MI to move to the right, MI of both hands simultaneously to move upwards, and resting (no MI) to move down. This paradigm is well established for 2D BCI control, and has been used in previous EEG-BCI studies to control devices like virtual cursors [9], [8], [29], [30], a quadcopter drone [21], and robotic arms [14], [15], [19].

The main feature of the BCI system proposed in this study is expanding the MI paradigm to allow users to interact with their environment instead of only moving in 2D. This is done by adding an output signal with two classes (i.e. Boolean: 0 or 1) to the MI paradigm, where one class can be used as the “off” or resting state (ex. 0), while the other class can be used as an “on” state, or “click” that denotes when the BCI system should perform an action. Some example potential uses of this additional output signal include clicking on objects with a virtual cursor (‘1’: click, ‘0’: don’t click), grabbing objects with a robotic arm (‘1’: grab / release, ‘0’: hold), or taking photographs with a drone (‘1’: take picture, ‘0’: rest).

In the proposed BCI system, subjects control the Boolean click signal by performing MI of one, or both, of their feet to output the “on” state (i.e. 1), and rest (no foot MI) to output the “off” state (i.e. 0). It is difficult to distinguish between left and right foot MI using EEG since the sensorimotor regions of both the left and right foot are anatomically located close together under the same EEG electrode. Because of this, MI of both feet is often covered by a single electrode in EEG BCI systems [31], [32]. Therefore, subjects were instructed that they could perform MI of either foot or both feet together, whichever they felt worked best to output a click for the tasks.

### D. Study Design

To evaluate the effectiveness of the proposed BCI system, subjects completed up to 7 sessions of BCI control featuring three different tasks. An illustration of each BCI task is shown in Fig. 2A and a graphical overview of the study design is included in Fig. 2B. Each BCI session lasted around 2 hours and each session for the same subject was completed on different days.

**Fig. 2.**
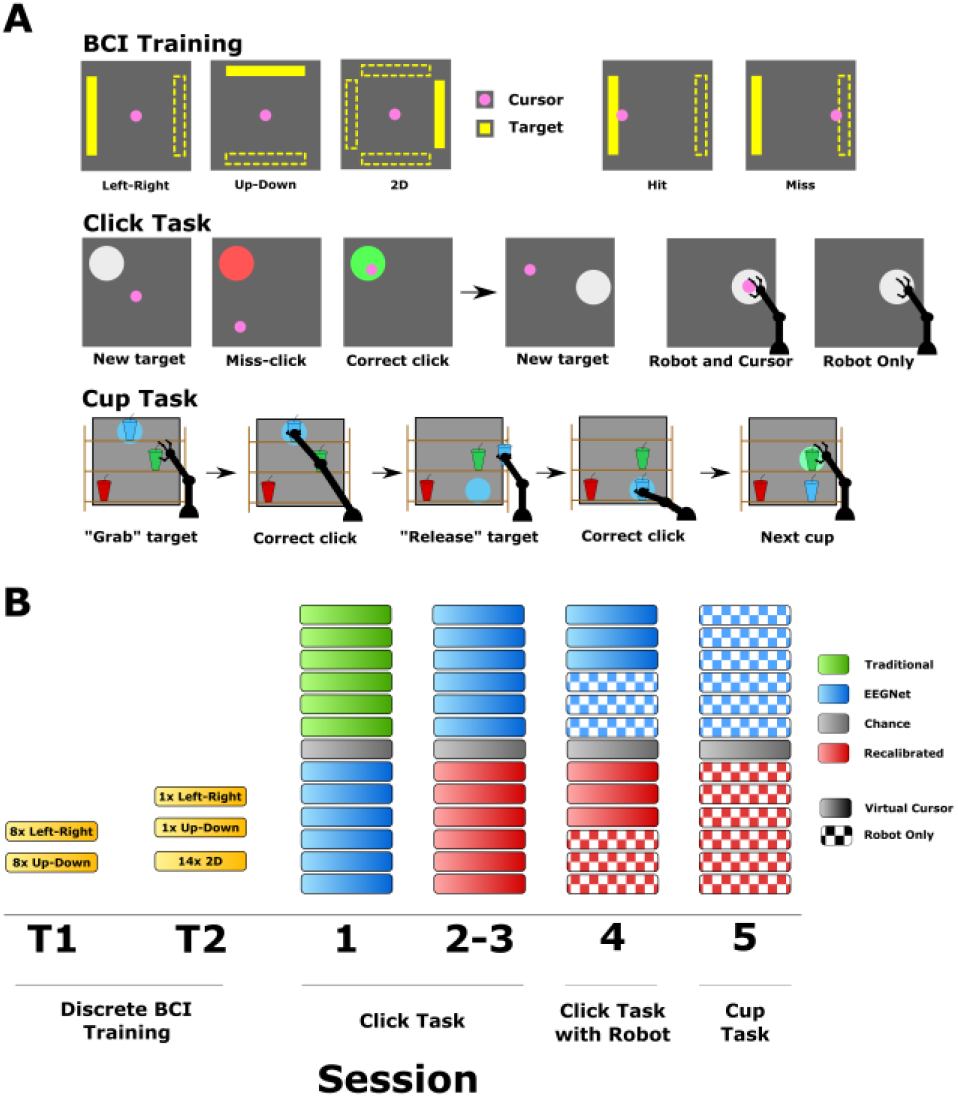
BCI Tasks and Study Design. A) Illustrations of the BCI tasks used in this study. During BCI training, a center-out cursor task was performed where targets were located on either the left and right, top and bottom, or all four sides of the workspace, depending on the task. In the click task, the subject attempted to move their cursor to the target area (light gray) and perform a click. If they clicked outside of the target area, the target would change to a red color to provide feedback to the subject. If they successfully clicked in the target area, it would turn green and pick a new random target location. In the cup task, three cups were arranged on a set of three vertical shelves. A random cup was chosen to start, and a matching-colored target was displayed on the screen behind the cup. The subjects attempted to move to the target cup and perform a click. On a successful click, the robotic arm would move forward and grab the cup, and a new random target was selected. On the next successful click, the robotic arm would move forward and place the cup on the shelf. After placing the cup, a new random cup was selected to continue the trial. B) An overview of the study design. Subjects first completed two sessions of BCI training. Sessions 1-3 were done with the click task and session 4 consisted of the click task using the robotic arm for feedback. In the final session, subjects performed the cup task.

The first two sessions consisted of BCI training using a traditional cursor movement task with discrete trials. The goal of these first two sessions was to provide new subjects with preliminary BCI experience and time to get acquainted with using the system before moving on to the main experiment. For subjects with prior BCI experience, these sessions also served to allow the subjects to refamiliarize themselves with the BCI system. The task used in the training sessions consisted of moving a virtual cursor (a pink circle) to collide with the virtual target (a yellow bar) that appeared on either the left, right, top, or bottom of the screen. Subjects were instructed to move the cursor by performing MI using the 2D MI paradigm described above (i.e. left-hand MI to move the cursor to the left). Each trial could end in one of three outcomes: a “hit” if the subject successfully moved the cursor to collide with the target, a “miss” if the cursor collided with one of the other sides of the screen, or an “abort” if no target was hit within 6 seconds.

The performance metric in this task is called Percent Valid Correct (PVC), and is calculated as the number of hit trials, divided by the number of hit and miss trials (excluding abort trials). The chance-level, or performance that can be expected by picking a random target, is 50% in 1D tasks and 25% in 2D. Two subjects were excluded from the remainder of the study since they were unable to obtain an average PVC above chance level in any of the training tasks in the first two sessions. This indicates that they were unable to obtain meaningful control of the BCI system, which is expected to occur with around 15-30% of all MI-BCI users [33].

After the initial BCI training, the first three sessions of the main study (Sessions 1-3) consisted of the virtual click task. This task used two BCI control signals, one to move a virtual cursor in two dimensions and another Boolean (i.e. 0 or 1) signal to output a “click”. The subjects’ goal in this task was to use the proposed MI paradigm to move the virtual cursor (a white circle) towards a larger, randomly placed target area (gray circle). Once inside of the target area, subjects attempted to click by performing foot MI. If a subject clicked while inside of the target area, a “hit” would be recorded, the target would turn green, and a new random target location would be chosen. If a subject clicked outside of the target area, a “miss” would be recorded, and the target would turn red to provide feedback to the subject. A two second “cooldown” period was applied after each click, whether hit or miss, before another click could be sent. This cooldown period was employed to prevent the decoder from clicking rapidly before subjects could react.

Subjects performed the same click task described above in Session 4, but also used a robotic arm for feedback. In the first three runs, the subjects moved both a virtual cursor and robotic arm simultaneously to perform the task. In the following three runs, the virtual cursor was made invisible so that the subjects could only use the robotic arm for feedback. Since previous works have shown that controlling a physical device may be more difficult than a virtual cursor [15], [19], the goal of this session was to introduce subjects to robotic arm feedback and gradually ramp up the difficulty of the tasks.

In the final session of the study, subjects attempted to control a robotic arm to pick up, move, and drop off three metal cups on a set of three vertical shelves placed in front of the screen (Fig. 1A). In this cup task, subjects used the same MI paradigm to move the robotic arm in 2D, then clicked using foot MI once the robotic arm was in the target area. At the beginning of each run, a colored cup (orange, green, or blue) was placed in the middle of each of the three shelves. One of the three cups was chosen at random to be the first target, and a colored target circle appeared on the screen behind that cup.

The subject’s first goal in this task was to move the robotic arm to that cup / target area, then use foot MI to send a click. On a successful click inside the target area, the robotic arm moved forward and grabbed the cup from the shelf. A new target area was then chosen and shown on the screen as a circle that matched the color of the cup. This new target area was chosen at random and could be on any of the three shelves (at a minimum distance from the other two targets to avoid collisions). The subject then attempted to use MI to move the robotic arm to the new target area and click using foot MI. If the subject was able to successfully click inside of the second target area, the robotic arm then moved forward and placed the cup on the shelf in the new location. Next, one of the other two cups was chosen at random to be the next target and the task continued. Each run of the cup task consisted of five 60 second trials, with a short 2 second break between trials while the robotic arm was reset back to the center of the workspace. The subjects’ goal in this task was to move as many cups as possible in the 5-minute run, and the positions of the cups were reset before each run.

Each session of the click or cup task consisted of 12 runs, where a run consisted of five 60 second trials with a small break between each run. After the first 6 runs in each session, an extra run was performed called the “chance” run. In this run, the screen was turned off and the subject was instructed to maintain a resting state without performing any MI. During this time, the BCI task was run in the background using the same decoder as the first six runs. The goal of this “chance” run was to estimate the baseline performance that the BCI system would achieve at random, without any intentional control from the subject. This performance is often called the chance level in other BCI tasks, since it is the performance that the system can achieve by random chance, and is useful for determining if the subjects can obtain meaningful control of the BCI system. After the chance run, the screen was turned back on, and the remaining six runs were completed as normal.

### E. DL-based BCI Decoding

The main decoder used in this study was a DL-based model adapted from the EEGNet architecture [34]. Supplementary Fig. 1A shows an overview of the DL decoding process. In this study, we used two different DL models with nearly identical architectures for each experiment: one to control the movement of the BCI system, and another to output the “click” signal. The movement DL model was trained to perform regression and output two continuous values, one for the horizontal velocity and another for the vertical velocity. The click model was trained to classify the EEG signals into a two-class output signal (Boolean), either a 0 for “don’t click” or a 1 for “click”. The DL models were subject-specific, meaning that each model was trained using only one subject’s BCI data and was to be used only by that specific subject. Details on the DL training process and computer hardware are provided in the Supplementary Materials.

Previous work has shown some preliminary evidence that recalibrating, or fine-tuning, the DL models during a BCI session may help to improve performance in continuous control tasks [30]. The hypothesis for this method is that fine-tuning on data from the current session may help the model overcome inter-session variability in the EEG and MI signals. To further investigate this technique, the DL models in each session were recalibrated halfway through the session by fine-tuning the models using the first six runs of data from the current session. The fine-tuning DL training was performed during the chance level runs by uploading the six runs of data to an external GPU server, fine-tuning the model, and downloading the resulting model weights. This process typically took around 5 minutes and was performed during the chance level runs in each session. As shown in Supplementary Fig. 1B, for the recalibration process, 5 runs were used for training, 1 run for validation, and the 6 runs in the second half of the session were used for testing.

### F. Statistical Analysis

Statistical testing was conducted using one-tailed, Wilcoxon-Signed rank tests using custom R scripts. The group-level results shown in Fig. 3 use a paired test, while subject-level results shown in Fig. 5 do not use a paired test. P-values were adjusted for multiple comparisons using the Holm method [35] unless noted otherwise. Significance is shown using stars with the following codes: n.s.: p>0.05, *: p<0.05, **: p<0.01, ***: p<0.001. Exact p-values were also shown if the value was above, but close to 0.05. Correlation values shown in Supplementary Fig. 2 were calculated as Pearson correlation coefficients. Boxplots were drawn in the style of Tukey, where outliers beyond 1.5 times the inter-quartile range are shown as individual points.

**Fig. 3.**
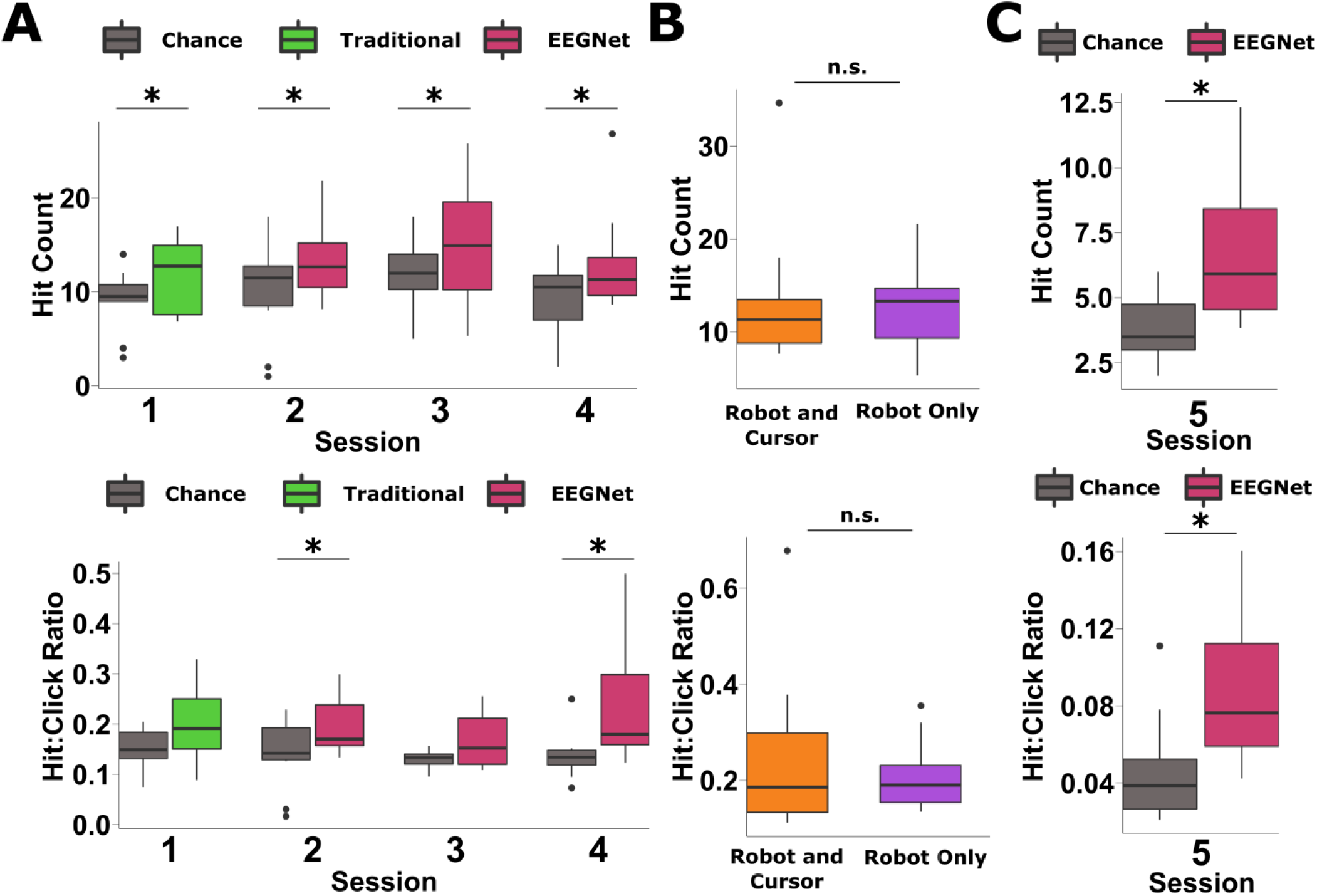
Group Level Results. A) Group level BCI performance across the first four sessions (after BCI training) in both hits per run (top) and hit-to-click ratio (bottom). Subjects performed the click task in each of these sessions. The traditional decoder was used in the first session and DL-based decoding in each session after. B) Performance in Session 4 with different feedback methods. The orange color is from runs that used both a virtual cursor and the robotic arm for feedback, while purple is from runs with only the robotic arm. C) Group level performance in Session 5, in which subjects performed the cup task. The top panel shows performance in the total number of hits per run, the bottom shows the hit-to-click ratio. Statistical testing was performed with a paired, one-tailed Wilcoxon-Signed Rank test. (n.s.: not significant, *: p<0.05)

**Fig. 4.**
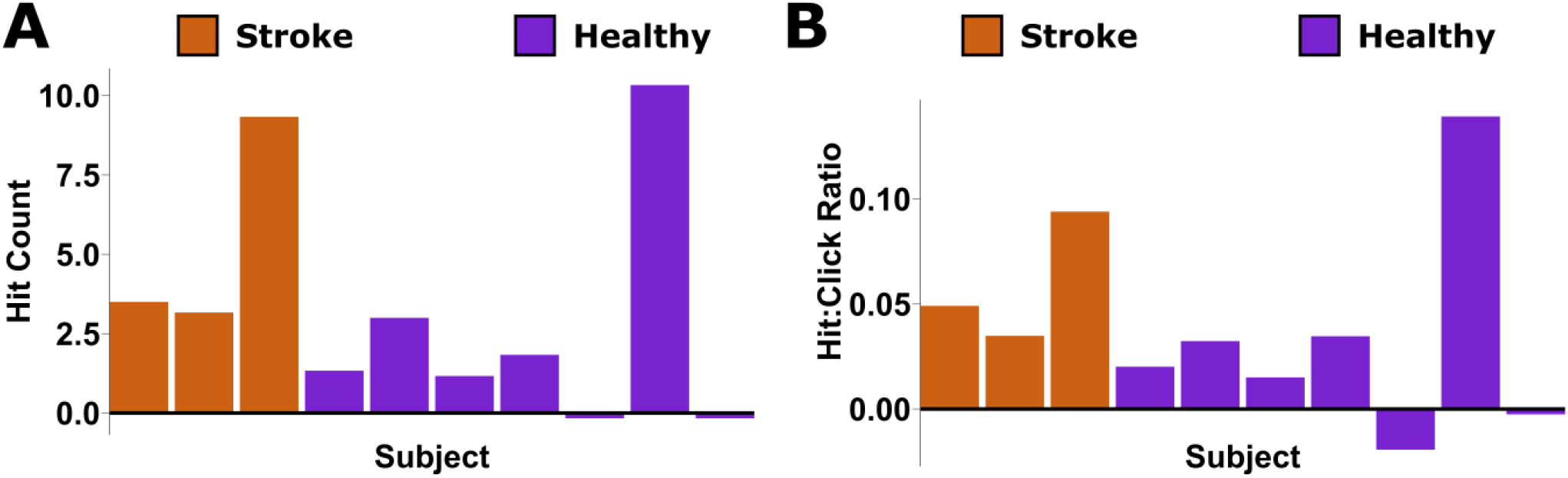
Subject Level Results. A) Subject-level performance for the cup task session (Session 5) shown as the total number of hits per run adjusted by chance level. To calculate this metric, each subject’s chance level performance for a specific session was subtracted from that subject’s average BCI performance in the same session to show how they performed compared to the chance level for their specific DL decoder. Healthy subjects are shown in purple while stroke-affected subjects are colored orange. B) The hit-to-click ratios for the same data.

**Fig. 5.**
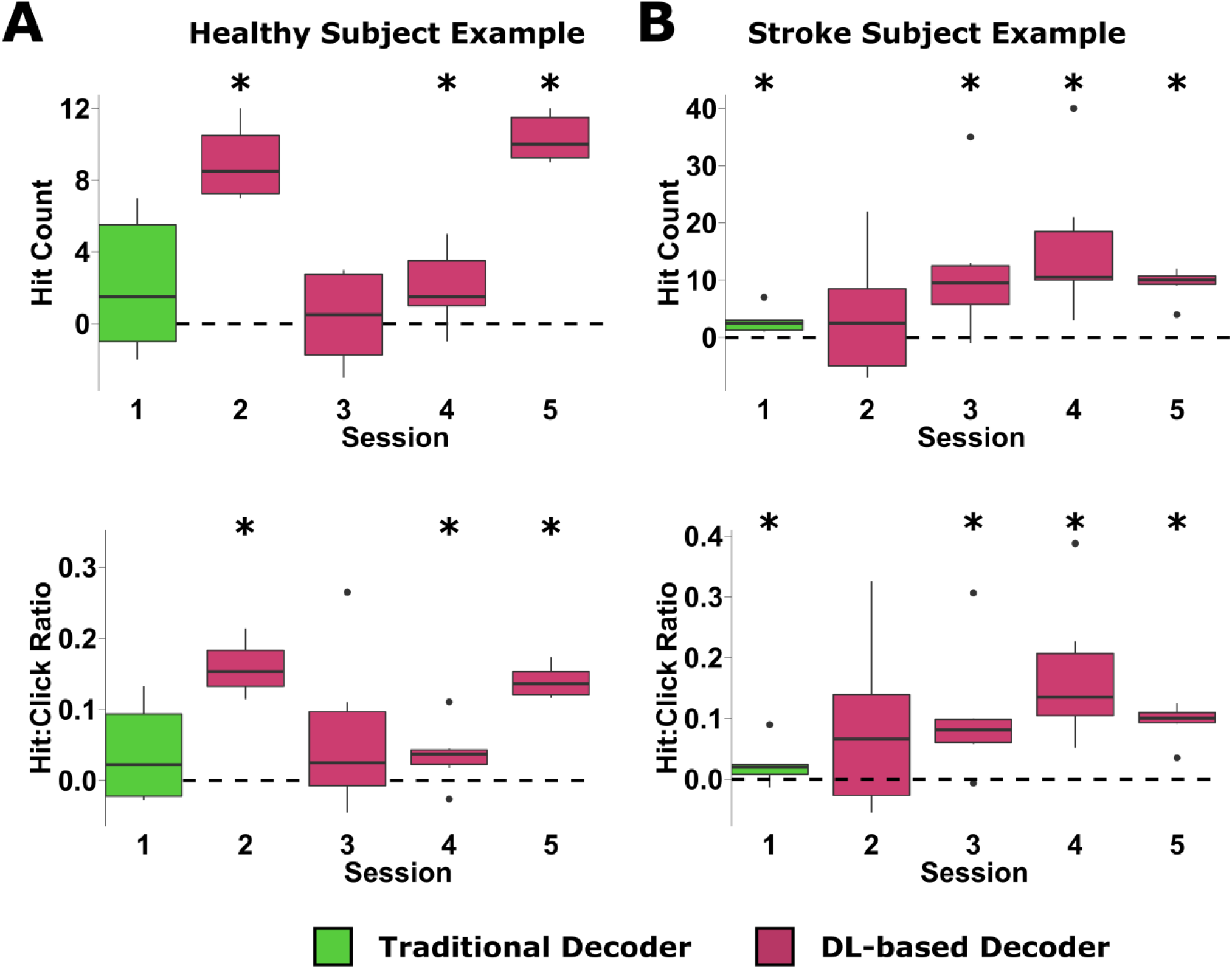
Session-level results for example subjects. A) The total hits per run (top) and hit-to-click ratio (bottom) performance for a high-performing healthy subject across all 5 sessions. Similar to the results shown in Figure 4, the results have been adjusted to this subject’s chance level by subtracting the chance level performance for each session from the average BCI performance for that session. B) The total hits per run (top) and hit-to-click ratio (bottom) for a high-performing stroke affected subject). Statistical testing was performed with a one-tailed Wilcoxon Signed Rank test. The significance values for these tests are shown without adjusting for multiple comparisons due to the low sample size when comparing data within a specific subject and session and each session being an independent analysis. Adjusting using the Holm method results in all tests being not significant (*: p<0.05).

## III. Results

### A. Performance Metrics

Two different BCI tasks were investigated in this study: the virtual click task and the cup task. In both paradigms, the subjects’ goal was to move their cursor (or robotic arm) to a target area using the 2D motor imagery paradigm, then send a “click” by performing foot MI. Trials continued after subjects clicked on a target by picking a new target location, and subjects were instructed to try to click on as many targets as possible during a 5-minute run.

Due to the unique design of these tasks, compared to previously studied BCI task paradigms, new methods to record subjects’ performance were required. Standard center-out cursor tasks have only one target per trial, so performance can be measured by calculating a trial-based accuracy, such as PVC in the training sessions. In contrast, the trials in this study continue after a target has been hit and do not result in a single hit-or-miss outcome, so PVC is not applicable.

The continuous pursuit paradigm, which the click tasks have been derived from, uses the average distance between the cursor and target as the performance metric. Here, a lower average distance indicates that the cursor is closer to the target, indicating better performance. This performance metric is also somewhat relevant for the continuous moving portion of the click tasks, but does not measure the clicking component of the task.

To effectively measure both the moving and clicking components of the proposed click tasks, new performance metrics needed to be developed. Since the subjects’ goal in both tasks was to click on as many targets as possible in each 5-minute run, the primary metric we observed was simply the number of targets hit in each run (number of hits or hit count). Since a successful hit requires both moving to the target area and clicking inside of it, this performance metric captures both the moving and clicking aspects of the task. One potential issue with the raw number of hits is that it does not provide any information about the accuracy of the “click” decoder. For example, if the “click” decoder constantly outputs a positive click signal, a subject may get a higher number of hits (since they only have to move to the target area), but will also have a very poor click accuracy (many of the clicks were outside of the target area). Examples of this concept in the data are shown in Supplementary Fig. 2 Panel A. There is a positive correlation between the total number of clicks and successful hits for both the chance level trials (r=0.41) and BCI trials (r=0.35). This indicates that a decoder that outputs more clicks will have more successful hits, but potentially at the cost of click accuracy.

To counter this, we also looked at the ratio of successful clicks (hits) compared to the total number of clicks (hits + misses). This hit-to-click ratio will be higher when more of the clicks are done inside of the target area, and lower when the decoder outputs clicks outside of the target area. Panel B of Supplementary Fig. 2 shows a scatter plot of the total number of clicks per run vs. the Hit-to-Click ratio. Here there is a negative correlation in both the change level runs (r=-0.3) and the BCI decoders (r=-0.39), indicating that blindly increasing the number of clicks may result in more hits, but can also lower the accuracy. Therefore, to fully characterize the subjects’ performance in the click tasks, it is important to look at both the number of successful hits that the subjects were able to achieve, as well as the click accuracy through the hit-to-click ratio.

### B. Group-Level Results

The group-level results are presented in Fig. 3 using both the total number of hits per run and the hit-to-click ratio as performance metrics. In Sessions 1-3 (session numbers do not include BCI training), the subjects performed the click task with a virtual cursor and with a robotic arm for feedback in Session 4. In the first session, a traditional (non-DL) BCI decoder was used to collect the initial training data for DL-based methods. In the click task sessions (Fig. 3A), the subjects were able to achieve a higher performance than chance level using both performance metrics, showing that, overall, subjects were able to use the proposed BCI system to complete the click task. There was, however, a wide range of performance with some subjects able to complete the task well, while others performed around chance level, indicating that they were unable to obtain meaningful control of the BCI system.

Previous studies using MI-based BCIs have shown some evidence that controlling a physical device, like a robotic arm, may be more challenging than a virtual cursor and can lead to slightly lower performance [15], [19]. To help transition subjects from the earlier sessions which used only a virtual cursor, to the later sessions using only a robotic arm for feedback, Session 4 consisted of runs using both the virtual cursor and robotic arm together, as well as some runs using only the robotic arm. Comparing the results of both feedback types (Fig. 3B), we found that using the robotic arm with and without the virtual cursor led to similar performances overall.

In the final session, the subjects performed the cup task, which consisted of using the BCI system to move physical cups around a vertical shelf with a robotic arm. At the group level (Fig. 3C), subjects achieved significantly better performance than chance level with both performance metrics. This shows that, overall, the subject group was able to complete this complex task to move physical objects using two different MI paradigms simultaneously.

The differences in performance between Sessions 1-4 and Session 5 (Figs. 3A and 3C) may be primarily due to logistic differences between the two tasks. The click task used a larger target than the cup task (1:0.58 diameter of the circles) to make the task easier for subjects while they gained experience with the BCI system. In addition, there was a down-time after each successful hit during the cup task while the robotic arm moved forward to grab or place the cup. This delay limited the maximum number of hits in the cup task to 7 per 1 minute trial (35 per run), while there was no delay in the click task except for the 2 second click cooldown, meaning subjects could get up to a maximum of 30 hits per trial. These two differences lowered the expected performance numbers in the cup task, due to the increased difficulty and decreased maximum possible performance in the task.

### C. Decoder Performance Variability

The proposed click tasks require the subject to complete two sub-tasks: moving to the target area, and sending a click once inside the target. One of the consequences of this paradigm is that the performance is reliant upon how often the click decoder sends a click. If a decoder is more likely to send clicks, whether intended by the subject or not, it will be more likely to get a higher number of hits and potentially lower hit-to-click ratios (Supplementary Fig. 2). Conversely, decoders that do not click as often will have lower hit counts. The results in Supplementary Fig. 2 show that this is the case for runs where the subject intentionally controls the BCI system, as well as in the chance level runs. Because of this phenomenon, there is a large variation in both BCI and chance level performance across subjects, and even across sessions, due to differences in how likely the decoders are to output a click signal.

Supplementary Fig. 3 shows the subject-level performances in both the BCI decoder and chance-level runs for a single session. These plots show that there is a large variation in performance in both the chance-level and decoder runs across subjects. And even though a subject may perform substantially better than the chance-level obtained with their decoder, the overall performance may be lower than the chance-level obtained with a different decoder. One primary reason for this is that some subjects’ click decoders may have optimized to send out many more clicks than others, resulting in vastly different baseline (chance-level) performances. For example, a decoder that outputs the maximum 30 clicks per trial is much more likely to get hits compared to a decoder that only outputs one click per trial, regardless of how well the subject can control the BCI movement.

To account for this variability, the subject-level results were adjusted for each decoder’s baseline performance by subtracting the chance-level results from a specific subject and session from the BCI performance obtained by the same subject and session. This allows us to compare the BCI performance of a subject using the exact same decoder when they are attempting BCI control vs. when they are at rest (random chance level).

The results of this adjustment then show how well each subject did in each session, relative to the chance-level obtained by that specific decoder. A value of 0 indicates that the subject obtained the same performance as the chance-level run. Positive values indicate that the subject outperformed the chance-level and were able to control the BCI.

## D. Subject-Level Results

The chance-level adjusted results for the cup task session (Session 5) are shown in Fig. 4. These results show the large range of performance variations across subjects. For both healthy subjects and stroke survivor groups, most subjects achieved performance above chance-level, indicating that they were able to perform the complex cup task with some control. However, other subjects were at or close to chance-level, showing that they were unable to obtain meaningful control of the BCI system, even after 5 sessions of BCI (including training). Finally, two subjects in particular achieved high performance in the cup task, one from each group.

Fig.5 shows the performance trends across each of the BCI sessions for the two high performing subjects, one healthy subject and one stroke survivor. In both examples, the subjects were able to achieve performances significantly higher than chance level in most sessions, in both tasks, and using both performance metrics. These results provide evidence that some subjects can do the complex tasks of virtual clicking and moving physical objects using the proposed MI paradigms of moving and clicking simultaneously.

### E. Recalibration

Previous work on the continuous pursuit BCI task has shown evidence that recalibrating, or fine-tuning the DL models mid-session, may help to overcome inter-session variations in the EEG signal and could improve performance. To investigate this, half-way through each session, during the chance-level runs, the DL models were recalibrated by fine-tuning them using the data from the first half of the session. The group-level results from the recalibration runs are shown in Supplementary Fig. 4 next to the results from the original models. In addition to the total number of hits per run and the hit-to-click ratio performance, the total number of clicks per run is also shown in the third panel.

The results show that the recalibrated models achieved a greater number of both hits and clicks compared to the original DL models in sessions 1, 2, and 4, while maintaining similar hit-to-click ratios. In sessions 3 and 5, recalibration achieved similar performance to the original models. Although these results suggest that recalibration may offer a modest improvement in performance, this improvement was not substantial enough to be statistically significant. In addition, it appears that the recalibration may not always be beneficial, as seen in sessions 3 and 5, but even in these cases it appears to offer similar performance.

## VI. Discussion

The goal of this study was to investigate the feasibility of expanding the degrees of freedom of EEG-based MI BCIs by adding an additional control signal. To this end, we have proposed a new motor imagery (MI) paradigm that include two simultaneous decoders: one to control 2D movement, and another to output a Boolean *click* signal, mimicking a computer mouse. To evaluate this system, we designed a virtual click task in which subjects moved in 2D to a randomly placed target area and initiated a click using volitional MI signals. We found that subjects were able to control the BCI in this complex task above chance-level, and that a few high-performing subjects could achieve substantial control in the task. To demonstrate other applications of this paradigm, we also designed an additional task in which subjects used an assistive robotic arm to grab and place physical cups around three vertical shelves. We found that a few of the subjects were able to achieve high performance in this complex task, and could move up to an average of 7 cups in 5 minutes (13.5 hits) using the BCI system. Adding the click output provides BCI users with an additional way to interact with the environment and the results from these experiments suggest that MI-based BCIs may be useful for complex applications like controlling assistive robotic devices.

### A. Behavioral Results

The group level results in Fig. 3 show that, overall, the subjects reached a performance significantly greater than chance-level in using both metrics in almost all sessions. This indicates that the subject cohort was able to obtain meaningful control over the BCI system, although the group-level performance was still limited. One reason for the relatively low performance is likely due to the increased complexity of the task, which is substantially more difficult than previous MI BCI tasks that only require 1D or 2D movement. Another reason may also be due to the inherent variability in human subject BCI performance. It is well established that there is a large range of BCI performance across human subjects [8], [36], and between 15-30% of users may not be able to produce strong enough MI features to control simple BCI tasks [33]. It may be the case that even fewer subjects are able to produce signals robust enough to perform the complex tasks proposed here.

The subject-level performances shown in Fig. 4 provide a more detailed view of the results. Most subjects across the sessions achieve a performance that is a modest improvement above chance level, showing that they can control the BCI, but not at a comfortable level. However, a few of the subjects achieved high performance in in the cup task, showing notable control over the BCI in this complex task. More work is needed in the EEG BCI field to investigate the differences in performance and MI features between subjects, and hopefully to find a method to improve the abilities of lower performing users.

### B. Stroke-Affected Subjects

One of the primary motivations for developing BCIs is to replace or restore motor functions for motor-impaired patients. To investigate whether stroke-affected users could use BCIs for complex tasks, like the click tasks proposed in this study, we recruited a cohort of human subjects who had previously suffered a stroke. Although the main application of BCIs may be for patients with significant motor-impairments, this initial exploration was not limited to stroke survivors with substantial motor impairments. The three stroke survivors who completed this study had initial motor impairment of an upper extremity at the time of their stroke, but all three had been recovered to near-normal functional levels by the time they were recruited for this study.

Interestingly, the subject-specific performance levels in Fig. 4 show similar overall performance levels between the healthy and stroke-affected cohorts. This initial exploration focused on testing the feasibility of using this BCI paradigm with a small number of stroke subjects, but provides preliminary evidence that complex MI BCIs may be useful for stroke-affected groups. Future work is needed to collect more data from a larger cohort in order to perform a robust statistical analysis, specifically in patients with active motor-impairments.

### C. Recalibration

In addition to inter-subject differences, there is also variation in the EEG signal and MI features in the same subject between different sessions. Previous work on DL for continuous pursuit BCI has shown that recalibrating, or fine-tuning, the DL models may help to improve performance by allowing the decoder to adapt to the inter-session feature differences. The results shown in Supplementary Fig. 4 show some supportive evidence of this, as the recalibration method achieved higher numbers of hits and clicks in some sessions while retaining similar hit-to-click ratios, but the improvement was not substantial enough to reach statistical significance. These results indicate that there may be merit in the recalibration method, but more research is needed on this topic, specifically on how to best implement the fine-tuning process.

### D. Challenges and Future Work

Adding the click output signal to the proposed MI paradigm introduces several new challenges to BCI development. One of the major challenges in this study was evaluating the performance of the BCI tasks. Unlike traditional BCI tasks like center-out cursor control, there can be multiple targets per trial so the task cannot be evaluated with a simple hit-or-miss accuracy. The main goal of the click tasks is for the subject to hit as many targets as possible within the time limit, so the primary performance metric we looked at was the number of hits per run. This metric provides an intuitive measure of how well the subject could control the BCI system with the proposed paradigm.

However, as shown in Supplementary Fig. 2, this performance metric is affected by how many “clicks” the decoder outputs. We also looked at the hit-to-click ratio to further analyze the performance, but this is also somewhat affected (in the opposite way) by the total number of clicks. In addition, these correlations also make it more difficult to evaluate chance level, as the number of clicks can have a large effect on the performance, even in chance level runs. Supplementary Fig. 3 shows that the chance level can be vastly different for each subject and session since different decoders are used, each with a different tendency to output clicks. We addressed this challenge by analyzing the results for each subject relative to the chance level obtained by the same decoder, but future studies will need to investigate this problem further or develop alternative performance metrics that are not dependent on the decoders’ tendency to output clicks.

The tendency of decoders to output clicks also presents an additional challenge for BCI design. In some applications, it may be better to have the system click more often if it also results in more hits and makes the system more responsive to the user. This may be the case in low-stakes situations where it is better to get more hits, even at the cost of more misses. However, there may be other cases where it is more important to make sure that all clicks are correct hits, even at the cost of obtaining relatively few hits. One example could be in applications that involve potentially dangerous scenarios where the user wants to be certain that the clicks are correct, such as moving a cup of hot coffee or sharp objects with the assistive robot. It is therefore important that BCI systems using a clicking paradigm like the one proposed here are designed and optimized around specific applications.

Another challenge in using DL for complex BCI tasks is obtaining the training data. The study design here used a method obtaining training data by having subjects directly perform the intended BCI task. In other words, subjects started doing the click task even in the first session (after BCI training). This allowed the subjects to get experience in the task at the same time as providing the initial training data for the DL models. However, an alternative training strategy could involve the subjects performing simpler tasks at first to obtain the initial DL training data, such as 1D center-out cursor control for foot MI, then moving on to the complex task once a model has been trained. The advantage of this alternative method could be to provide cleaner initial training data to the DL models from an easier task where subjects are more likely to perform well. However, two potential disadvantages of this method are that it may require more BCI training sessions to get to the evaluation tasks, and that features learned in the simpler task may not be the same as those produce by the subjects in the more complex tasks. This could make it difficult to transfer trained DL models between the two different tasks. Future studies could be done to investigate the optimal method of obtaining DL training data for complex BCI tasks such as these.

## V. Conclusion

Recent studies involving EEG BCIs have shown that these devices can be used to control robotic limbs in 2D and completing tasks involving selecting from a discrete set of actions. In this study, we propose an MI paradigm that extends 2D BCI control by adding an additional Boolean output to mimic the click of a computer mouse. We evaluated this paradigm in a virtual cursor task and a task controlling a robotic arm to move physical objects, showing that healthy and stroke-affected subjects were able to perform complex tasks. These results have larger implications as well. With robust performance, this BCI paradigm could be used to control a computer mouse to do everyday tasks on a PC or give motor-impaired patients more autonomy when interacting with their physical environment using assistive robotic devices. Overall, this study represents a step forward in the development of MI-based EEG BCIs and moving towards real-world or clinical applications of this technology.

## Data Availability

Experimental data produced from this study will be made publicly available online after the work is accepted for publication at a peer-reviewed journal.

## Author Contributions

Conceptualization & study design: D.F., B.H. Investigation: D.F., G.F.W., B.H. Formal analysis: D.F. Data acquisition: D.F., Y.Z. Writing of original manuscript: D.F. Manuscript revision: D.F., G.F.W., and B.H. Supervision: B.H. G.F.W. also facilitated stroke patient recruitment and provided expertise on human subject research with motor-impaired patients.

## References

[1] K. K. Ang et al., “A Randomized Controlled Trial of EEG-Based Motor Imagery Brain-Computer Interface Robotic Rehabilitation for Stroke,” Clin EEG Neurosci, vol. 46, no. 4, pp. 310–320, Oct. 2015, doi: 10.1177/1550059414522229.

[2] M. A. Cervera et al., “Brain-computer interfaces for post-stroke motor rehabilitation: a meta-analysis,” Ann Clin Transl Neurol, vol. 5, no. 5, pp. 651–663, Mar. 2018, doi: 10.1002/acn3.544.

[3] B. He, B. Baxter, B. J. Edelman, C. C. Cline, and W. W. Ye, “Noninvasive Brain-Computer Interfaces Based on Sensorimotor Rhythms,” Proceedings of the IEEE, vol. 103, no. 6, pp. 907–925, Jun. 2015, doi: 10.1109/JPROC.2015.2407272.

[4] B. He, J. Yuan, J. Meng, and S. Gao, “Brain-Computer Interfaces,” in Neural Engineering, 3rd ed., Springer International Publishing, Cham, 2020, pp. 131–183.

[5] K. Värbu, N. Muhammad, and Y. Muhammad, “Past, Present, and Future of EEG-Based BCI Applications,” Sensors, vol. 22, no. 9, Art. no. 9, Jan. 2022, doi: 10.3390/s22093331.

[6] H. Yuan and B. He, “Brain–Computer Interfaces Using Sensorimotor Rhythms: Current State and Future Perspectives,” IEEE Transactions on Biomedical Engineering, vol. 61, no. 5, pp. 1425–1435, May 2014, doi: 10.1109/TBME.2014.2312397.

[7] J. R. Wolpaw and D. J. McFarland, “Control of a two-dimensional movement signal by a noninvasive brain-computer interface in humans,” Proceedings of the National Academy of Sciences, vol. 101, no. 51, pp. 17849–17854, Dec. 2004, doi: 10.1073/pnas.0403504101.

[8] J. R. Stieger, S. Engel, H. Jiang, C. C. Cline, M. J. Kreitzer, and B. He, “Mindfulness Improves Brain–Computer Interface Performance by Increasing Control Over Neural Activity in the Alpha Band,” Cereb Cortex, vol. 31, no. 1, pp. 426–438, Sep. 2020, doi: 10.1093/cercor/bhaa234.

[9] J. Meng, T. Streitz, N. Gulachek, D. Suma, and B. He, “Three-Dimensional Brain–Computer Interface Control Through Simultaneous Overt Spatial Attentional and Motor Imagery Tasks,” IEEE Transactions on Biomedical Engineering, vol. 65, no. 11, pp. 2417–2427, Nov. 2018, doi: 10.1109/TBME.2018.2872855.

[10] R. Leeb, D. Friedman, G.R. Müller-Putz, R. Scherer, M. Slater, and G. Pfurtscheller, “Self-Paced (Asynchronous) BCI Control of a Wheelchair in Virtual Environments: A Case Study with a Tetraplegic,” Computational Intelligence and Neuroscience, vol. 2007, p. e79642, Sep. 2007, doi: 10.1155/2007/79642.

[11] F. Galán et al., “A brain-actuated wheelchair: Asynchronous and non-invasive Brain–computer interfaces for continuous control of robots,” Clinical Neurophysiology, vol. 119, no. 9, pp. 2159–2169, Sep. 2008, doi: 10.1016/j.clinph.2008.06.001.

[12] Y. Li, J. Pan, F. Wang, and Z. Yu, “A Hybrid BCI System Combining P300 and SSVEP and Its Application to Wheelchair Control,” IEEE Transactions on Biomedical Engineering, vol. 60, no. 11, pp. 3156–3166, Nov. 2013, doi: 10.1109/TBME.2013.2270283.

[13] L. Tonin, R. Leeb, A. Sobolewski, and J. D. R. Millán, “An online EEG BCI based on covert visuospatial attention in absence of exogenous stimulation,” J. Neural Eng., vol. 10, no. 5, p. 056007, Oct. 2013, doi: 10.1088/1741-2560/10/5/056007.

[14] J. Meng, S. Zhang, A. Bekyo, J. Olsoe, B. Baxter, and B. He, “Noninvasive Electroencephalogram Based Control of a Robotic Arm for Reach and Grasp Tasks,” Sci Rep, vol. 6, no. 1, Art. no. 1, Dec. 2016, doi: 10.1038/srep38565.

[15] B. J. Edelman et al., “Noninvasive neuroimaging enhances continuous neural tracking for robotic device control,” Science Robotics, vol. 4, no. 31, p. eaaw6844, Jun. 2019, doi: 10.1126/scirobotics.aaw6844.

[16] J.-H. Jeong, K.-H. Shim, D.-J. Kim, and S.-W. Lee, “Brain-Controlled Robotic Arm System Based on Multi-Directional CNN-BiLSTM Network Using EEG Signals,” IEEE Transactions on Neural Systems and Rehabilitation Engineering, vol. 28, no. 5, pp. 1226–1238, May 2020, doi: 10.1109/TNSRE.2020.2981659.

[17] V. Mondini, R. J. Kobler, A. I. Sburlea, and G.R. Müller-Putz, “Continuous low-frequency EEG decoding of arm movement for closed-loop, natural control of a robotic arm,” J Neural Eng, vol. 17, no. 4, p. 046031, Aug. 2020, doi: 10.1088/1741-2552/aba6f7.

[18] R. Zhang et al., “NOIR: Neural Signal Operated Intelligent Robots for Everyday Activities,” in Proceedings of The 7th Conference on Robot Learning, J. Tan, M. Toussaint, and K. Darvish, Eds., in Proceedings of Machine Learning Research, vol. 229. PMLR, Nov. 2023, pp. 1737–1760. [Online]. Available: https://proceedings.mlr.press/v229/zhang23f.html

[19] D. Forenzo and B. He, “Online Robotic Arm Control with a Deep Learning-Based EEG BCI,” in 2024 10th IEEE RAS/EMBS International Conference for Biomedical Robotics and Biomechatronics (BioRob), Sep. 2024, pp. 832–837. doi: 10.1109/BioRob60516.2024.10719774.

[20] A. J. Doud, J. P. Lucas, M. T. Pisansky, and B. He, “Continuous Three-Dimensional Control of a Virtual Helicopter Using a Motor Imagery Based Brain-Computer Interface,” PLOS ONE, vol. 6, no. 10, p. e26322, Oct. 2011, doi: 10.1371/journal.pone.0026322.

[21] K. LaFleur, K. Cassady, A. Doud, K. Shades, E. Rogin, and B. He, “Quadcopter control in three-dimensional space using a noninvasive motor imagery based brain-computer interface,” J Neural Eng, vol. 10, no. 4, p. 10.1088/1741-2560/10/4/046003, Aug. 2013, doi: 10.1088/1741-2560/10/4/046003.

[22] B. J. Edelman et al., “Non-invasive Brain-Computer Interfaces: State of the Art and Trends,” IEEE Reviews in Biomedical Engineering, pp. 1–25, 2024, doi: 10.1109/RBME.2024.3449790.

[23] L. R. Hochberg et al., “Reach and grasp by people with tetraplegia using a neurally controlled robotic arm,” Nature, vol. 485, no. 7398, pp. 372–375, May 2012, doi: 10.1038/nature11076.

[24] J. L. Collinger et al., “High-performance neuroprosthetic control by an individual with tetraplegia,” Lancet, vol. 381, no. 9866, pp. 557–564, Feb. 2013, doi: 10.1016/S0140-6736(12)61816-9.

[25] B. Wodlinger, J. E. Downey, E. C. Tyler-Kabara, A. B. Schwartz, M. L. Boninger, and J. L. Collinger, “Ten-dimensional anthropomorphic arm control in a human brain-machine interface: difficulties, solutions, and limitations,” J Neural Eng, vol. 12, no. 1, p. 016011, Feb. 2015, doi: 10.1088/1741-2560/12/1/016011.

[26] S. N. Flesher et al., “A brain-computer interface that evokes tactile sensations improves robotic arm control,” Science, vol. 372, no. 6544, pp. 831–836, May 2021, doi: 10.1126/science.abd0380.

[27] A. B. Ajiboye et al., “Restoration of reaching and grasping movements through brain-controlled muscle stimulation in a person with tetraplegia: a proof-of-concept demonstration,” Lancet, vol. 389, no. 10081, pp. 1821–1830, May 2017, doi: 10.1016/S0140-6736(17)30601-3.

[28] G. Schalk, D. J. McFarland, T. Hinterberger, N. Birbaumer, and J. R. Wolpaw, “BCI2000: A General-Purpose Brain-Computer Interface (BCI) System,” IEEE Trans. Biomed. Eng., vol. 51, no. 6, pp. 1034–1043, Jun. 2004, doi: 10.1109/TBME.2004.827072.

[29] D. Forenzo, Y. Liu, J. Kim, Y. Ding, T. Yoon, and B. He, “Integrating Simultaneous Motor Imagery and Spatial Attention for EEG-BCI Control,” IEEE Trans Biomed Eng, vol. PP, Jul. 2023, doi: 10.1109/TBME.2023.3298957.

[30] D. Forenzo, H. Zhu, J. Shanahan, J. Lim, and B. He, “Continuous tracking using deep learning-based decoding for noninvasive brain–computer interface,” PNAS Nexus, vol. 3, no. 4, p. pgae145, Apr. 2024, doi: 10.1093/pnasnexus/pgae145.

[31] G. Pfurtscheller, C. Brunner, A. Schlögl, and F. H. Lopes da Silva, “Mu rhythm (de)synchronization and EEG single-trial classification of different motor imagery tasks,” NeuroImage, vol. 31, no. 1, pp. 153–159, May 2006, doi: 10.1016/j.neuroimage.2005.12.003.

[32] G.R. Müller-Putz, V. Kaiser, T. Solis-Escalante, and G. Pfurtscheller, “Fast set-up asynchronous brain-switch based on detection of foot motor imagery in 1-channel EEG,” Med Biol Eng Comput, vol. 48, no. 3, pp. 229–233, Mar. 2010, doi: 10.1007/s11517-009-0572-7.

[33] B. Blankertz et al., “Neurophysiological predictor of SMR-based BCI performance,” Neuroimage, vol. 51, no. 4, pp. 1303– 1309, Jul. 2010, doi: 10.1016/j.neuroimage.2010.03.022.

[34] V. J. Lawhern, A. J. Solon, N. R. Waytowich, S. M. Gordon, C. P. Hung, and B. J. Lance, “EEGNet: a compact convolutional neural network for EEG-based brain–computer interfaces,” J. Neural Eng., vol. 15, no. 5, p. 056013, Jul. 2018, doi: 10.1088/1741-2552/aace8c.

[35] S. Holm, “A Simple Sequentially Rejective Multiple Test Procedure,” Scandinavian Journal of Statistics, vol. 6, no. 2, pp. 65–70, 1979.

[36] S. Saha and M. Baumert, “Intra- and Inter-subject Variability in EEG-Based Sensorimotor Brain Computer Interface: A Review,” Front. Comput. Neurosci., vol. 13, Jan. 2020, doi: 10.3389/fncom.2019.00087.

