## Supplementary Materials for "Continuous Reaching and Grasping with a BCI Controlled Robotic Arm in Healthy and Stroke-Affected Individuals"

Of

### Supplementary Methods:

*A. DL Model Paradigm*

Two DL decoders were trained for every subject in every session. One model was used to control the 2D movement of the virtual cursor or robotic arm. This model performed regression and output two continuous signals, one for the horizontal velocity and one controlling the vertical velocity of the cursor or arm. The second DL model was used for the click signal. This model performed classification, classifying the signal into one of two classes: 0 or 1. In this way, the click model outputs a Boolean (0 or 1) every 40ms, creating a continuous Boolean signal.

The two DL decoders used almost identical model architectures, based on the EEGNet architecture, but the main differences between the two were the data-labelling method that was used for supervised learning and the performance metric used for validation. Illustrations of the labelling methods and performance metrics used for each type of model are provided in Supplementary Fig. 1C. First, the continuous EEG signal was cut into 1-second-long windows that overlapped every 40ms. Since new data packets sampled at 1 kHz were sent every 40ms during the online experiment, this allows the processing methods to be used in both online and offline analysis. For the DL model that controlled movement, the model was trained to predict a cursor velocity by using the label as the average vector between the cursor and target locations throughout the 1s window. In this way, the DL model learned to predict the optimal direction to move the cursor to get closer to the target. To validate this model, we used the angle between the predicted direction and the calculated label as the performance metric. A smaller angle indicated that the model was predicting close to the optimal direction, while a larger angle indicated worse performance. More details about training DL models for this continuous 2D movement paradigm can be found in previous work [30].

To train a DL model to predict the "click" signal, we used a Boolean label of either 0 for "don't click" or 1 for "click". If the cursor was inside the target area, then the subject should be trying to click by performing foot MI, so that window should be labelled with a 1. If the cursor was outside of the target area, then the subject was not supposed to be performing foot MI, and the data window should be labelled with a 0. To obtain these labels, the cursor positions throughout the 1 second window were compared to the current target area. If the cursor was inside of the target area for more than 50% of a particular data window, then the sample was labelled as class 1 for the click decoder, and as class 0 if not. Using this labelled method, the DL model could be trained to do a two-class classification task to produce the Boolean click signal. For validation, the accuracy of the model was calculated on samples labelled with each class separately to get a performance metric for "click" samples and another for the "don't click" samples. The total accuracy was obtained by weighing each class accuracy by the number of samples in each class, to avoid a bias towards one class due to large class imbalances.

The training data split is shown in Supplementary Fig. 1B. Each DL model was trained using all the data from a specific subject's previous BCI sessions. For each session, the 6$^{th}$ and 12$^{th}$ runs were used as a validation set while the rest were used for training, resulting in a 5:1 training-to-validation split. For the recalibration models, only 6 runs from the current session were used to fine-tune the model. Five runs were used for training and one (the 4$^{th}$ run) was used for validation. This method allowed us to use most of the runs for model fine-tuning, while also having a validation set.

*B. DL Training Details*

DL models were trained for 15 epochs using an Adam optimizer with a learning rate of 0.005. After the main training loop, the weights that achieved the best validation performance were kept as the trained model weights. For the movement model, the loss was calculated as the Mean Squared Error (MSE) between the x and y positions of both the label and prediction vectors. An additional KL-divergence term was added to the loss to try to coerce the model to make predictions following a Gaussian distribution with mean 0 and unit standard deviation. Training the movement models followed the same procedure as in [30], and more details can be found there. For the click decoders, the loss was calculated as the cross entropy between the prediction and label classes with weights corresponding to the fraction of each class among the training data.

DL training was performed on a custom-built GPU server (Exxact Corporation). This server had the following specifications: 2 24-core AMD EPYC 7413 processors, 512 GB of RAM, 7 NVIDIA RTX A5000 GPUs with 24 GB VRAM. Data was transferred to the server from the desktop PCs used to run the online experiments after each session. The DL models were trained remotely on this GPU server and trained model weights were downloaded back to the desktop PC to be used for the following session. For fine-tuning models mid-session (recalibration), the data were uploaded to the server at the start of the 5-minute chance-level runs and the training was run during the chance level trials. Upon completion, the weights were downloaded back to the desktop PC and the experiment continued with the recalibrated model weights.

BCI experiments were performed on a desktop workstation with the following specifications: Dell XPS 8930 with a 6-core Intel i7-8700, 64GB of RAM, and an NVIDIA GeForce GTX 1070 Ti.

*C. Traditional BCI Decoder*

One drawback of using the DL-based models for BCI decoding is that these methods typically require an initial dataset to train the models. Therefore, a different type of decoder was required for the first few BCI runs to collect training data. For the first half of the first BCI session (after training sessions), we used a simple traditional BCI decoder: the BCI2000 ARSignalProcessing module. This decoder is well established, has been used extensively for many previous MI BCI studies and is available from the default BCI2000 installation. Details about using the AR decoder for 2D movement with hand MI can be found in previous publications [8],[9],[15], and the same method was used for 2D cursor control in this study.

To produce the "click" signal, the band power of the upper-mu rhythm frequency band of the EEG signal in electrode Cz was extracted in a 3Hz bin centered around 12Hz. The band power was then multiplied by -1 so that a decrease in band power due to MI ERD would cause an increase in the output signal. This signal was then normalized (z-scored) over the previous 30 seconds so that the average signal was 0, ERD due to foot MI would cause a positive signal, and resting would cause a negative signal in the ideal case. This signal was then added to a rolling sum (every 40ms), and once if the sum ever reached above a threshold (default 20) so that only several positive values in a row would cause a “click”.

*D. Robotic Arm Control*

A Kinova JACO assistive robotic arm was used to provide feedback to subjects during some BCI tasks and to move physical objects during the cup task. The robotic arm was mounted on a table in front of subjects at a safe distance during experiments. Safety boundaries were set up to prevent the arm from moving too close to the subject or the television monitor used to display the virtual task. To control the robotic arm, the cursor positions from the BCI application were sent out of BCI2000 every 40ms over an UDP socket on the local machine (a feature included in BCI2000). A custom robotic arm controller, written in C++ using the JACO API, was run on a loop to read the most recent cursor positions from the UDP socket, translate them from the virtual task coordinates to physical units, send a command to the robotic arm to move to the designated physical location, and repeat. This method allowed the subject to control the robotic arm using the same BCI2000 system as with the virtual tasks.

During the cup task, the robotic arm was also required to grab and release physical objects onto a set of vertical shelves. To accomplish these maneuvers, additional information was sent from BCI2000 to the robotic arm controller during this task. Upon a successful click in the target area, a positive Boolean “click” signal was sent to indicate that the arm would need to perform a grasp or release. At this time, BCI2000 also sent the exact location of the current target and another Boolean indicating whether the arm should perform a grab or a release. The arm was the programmed to move to the exact target location, move forward towards the screen a specific distance, either close or open the gripper fingers (depending on whether it was performing a grab or a release), and then move away from the screen the same distance to return to the original workspace plane. This method of hardcoding the movements allowed the robotic arm to avoid collisions with the vertical shelves and television monitor and could be used to pick up and place cups multiple times without needing human operator intervention.

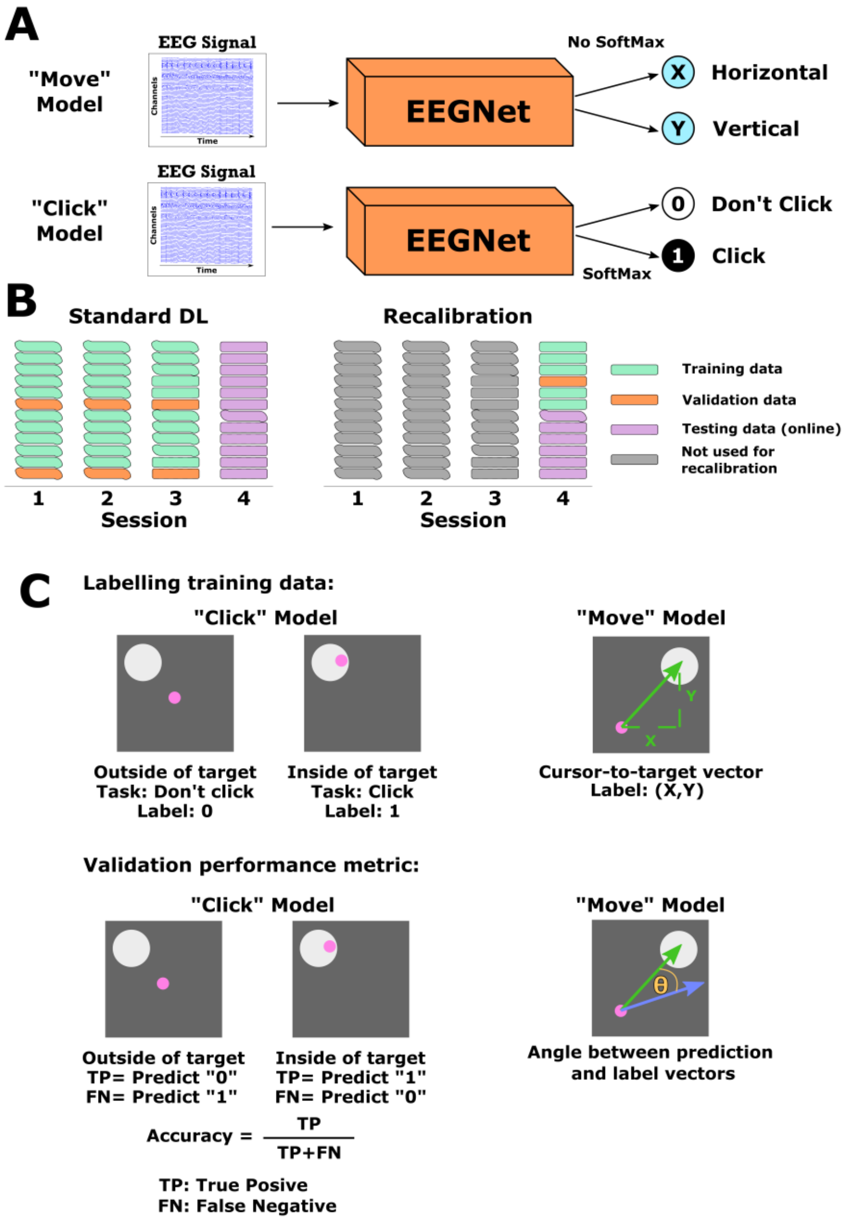


Supplementary Figure 1. Deep learning-based BCI decoding. A) An overview of the DL pipeline for BCI decoding. The EEG signal is given as an input to the DL models as a 2D matrix where each row corresponds to one EEG channel and each column is a single time point. The DL model, a modified version of EEGNet in this case, then processes the EEG signal through several steps called layers. Two different DL models were trained for each session: one to control 2D movement, and other to provide the “click” Boolean output signal. B) The training data split used in the study. For standard DL models, all of the sessions for a specific subject were used to train the models for that subject’s upcoming session. The 6$^{th}$ and 12$^{th}$ runs were used for validation while the rest were used as training data. For recalibration models, the model starts with pre-trained weights for a session and only the six runs from the current session are used to fine-tune the model. Five runs were used for training and a single run for validation. C) Data labelling and validation. Data was labelled for supervised learning in two different ways. For the click model, a data window got a label of “0” or “don’t click” if the cursor is outside the target area, and a “1” or “click” if it was inside the target area. For the movement model, the vector from the cursor to the target was used as the ground truth label, since this is the ideal direction that the subject should try to move. To validate the performance of each model, the accuracy of the 2-class classification task was used for the click model, while the average angle between the prediction and label was used for the movement model.

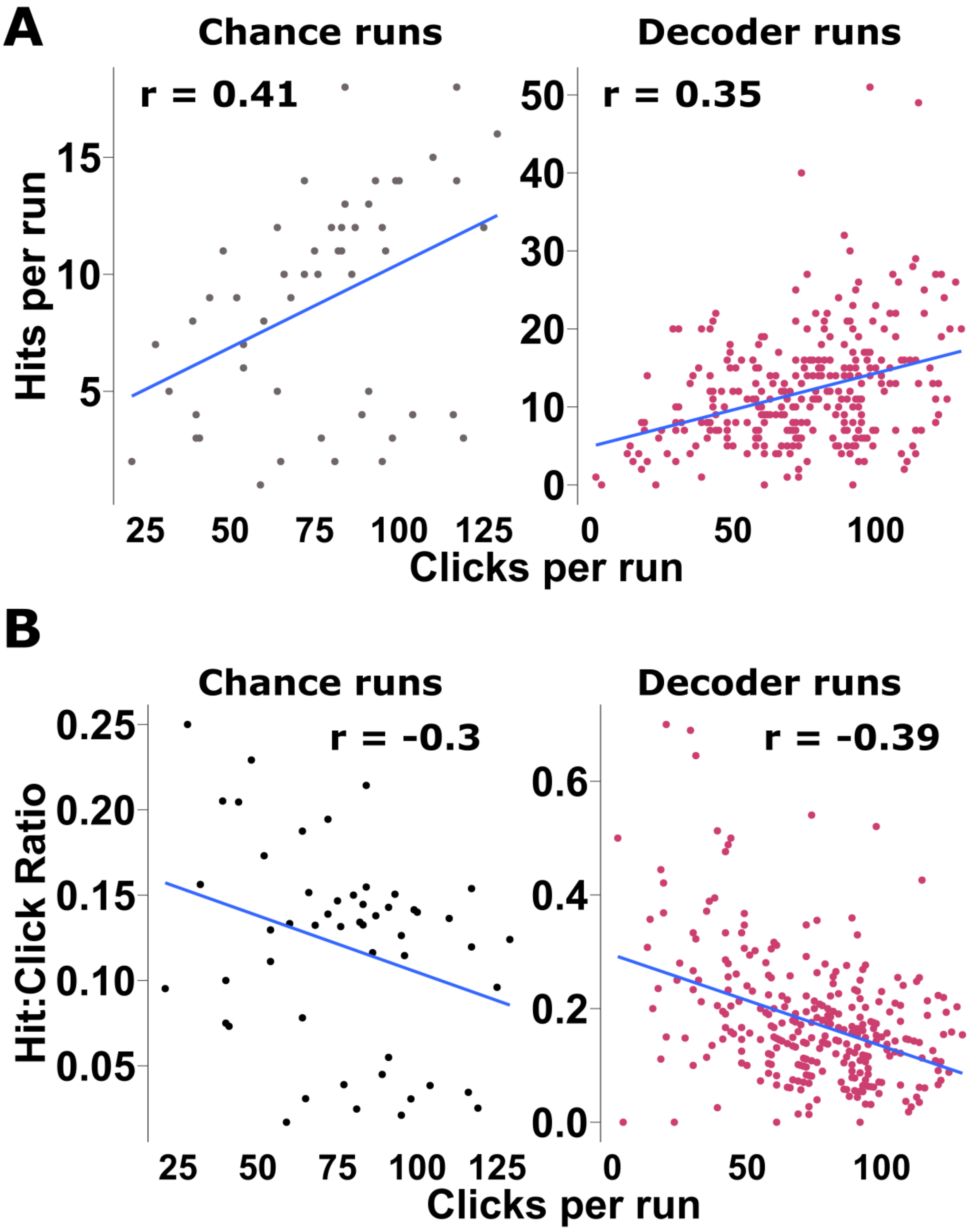


Supplementary Figure 2. Performance metrics and trends. A) Scatter plots comparing the number of successful clicks per run (hits) vs. the total number of clicks per run for both chance level runs (left) and runs with BCI decoding (right). The correlation is shown above the plot, and the trend line is shown in blue. B) Scatterplots comparing the hit-to-click ratio vs. the total number of clicks per run. The hit-to-click ratio is calculated as the number of successful clicks (hits) in a run divided by the total number of clicks in a run.

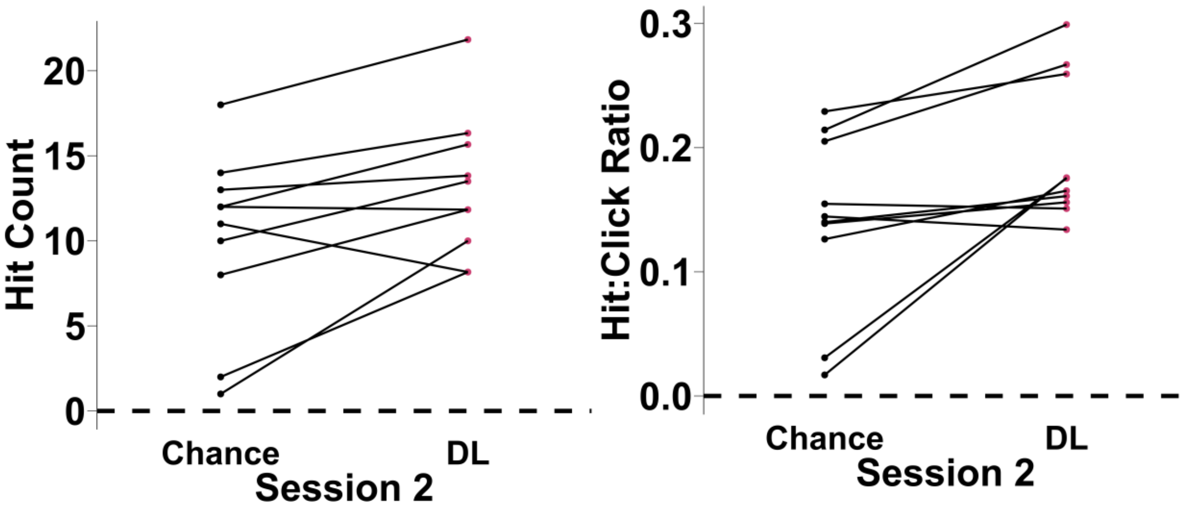


Supplementary Figure 3. Individual subject performance in BCI and chance-level runs. Each subject's performance using the DL-based decoder in BCI runs is shown in red on the right column and connected to the chance-level performance using that same DL-based decoder on the left in black. These connections show that there is a wide range of performances for each decoder in both BCI and chance-level runs. Even though some subjects outperformed the chance-level for their decoder, their performance may be lower than chance level for other decoders that outputted more clicks.

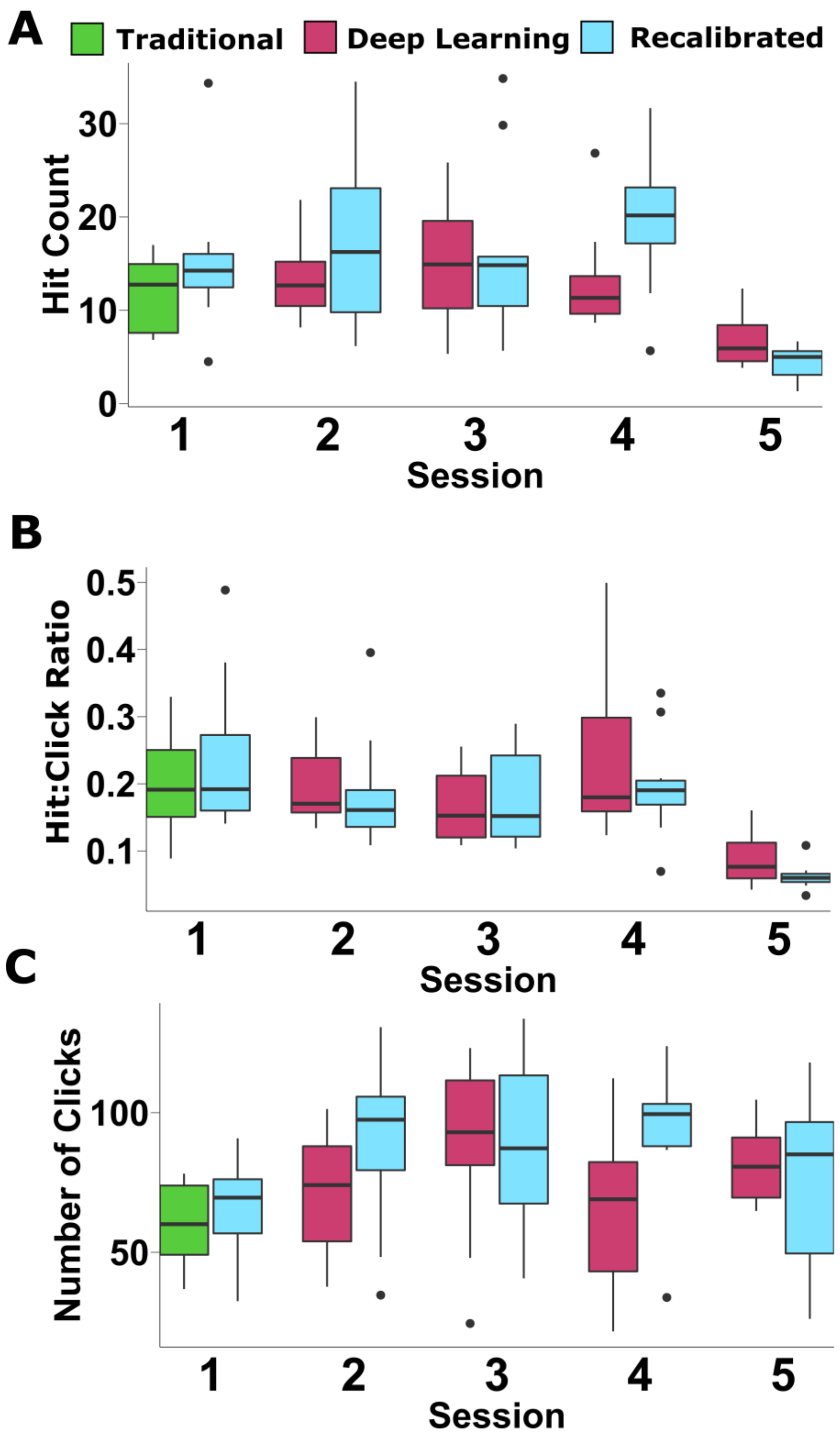


Supplementary Figure 4. Group-level performance for of recalibrated models. In each panel, the original models are shown in green (traditional) or red (DL), while recalibrated models are in light blue. The results are shown for Hit count (A), Hit-to-click ratio (B), and total number of clicks (C). Although not statistically significant after adjusting for multiple comparisons, recalibration resulted in more hits and overall clicks with similar hit-to-click ratios.
